# Prediction of adolescent suicide attempt by integrating clinical, neurocognitive and geocoded neighborhood environment data

**DOI:** 10.1101/2022.06.13.22276356

**Authors:** Elina Visoki, Tyler M. Moore, Ruben C. Gur, Victor M. Ruiz, Joel A. Fein, Tami D. Benton, Raquel E. Gur, Fuchiang R. Tsui, Ran Barzilay

**Author notes:** **Corresponding Author:** Ran Barzilay, MD, PhD, 10th floor, Gates Pavilion, Hospital of the University of Pennsylvania, 34th and Spruce Street, Philadelphia, PA 19104. **Statistical expert:** Tyler M. Moore, PhD.

## Abstract

**Objective:** The authors used multimodal data collected during pre/early-adolescence in research settings to predict self-report of past suicide attempt (SA) by mid-late adolescence in pediatric care settings. The study further aimed to determine types of data that contribute to the prediction; and to test generalizability of the prediction in an independent sample.

**Methods:** The authors applied machine learning methods on clinical, neurocognitive and geocoded neighborhood environmental data from the Philadelphia Neurodevelopmental Cohort study (PNC, Mean age 11.1, SD=2.2, 52.3% female and 51.4% Black participants) to predict SA reported ∼5 years later in a community pediatric clinic (n=922, 5.3% SA) or emergency department (n=497, 8.2% SA). The authors compared prediction performance when using all data versus individual data types, then used feature selection algorithms (Lasso, Relief and Random Forest) to identify important predictors and compared performance of models relying on feature subsets.

**Results:** In the training dataset, SA prediction was good, reaching AUC=0.75, sensitivity/specificity 0.76/0.77 when relying on feature subsets identified using feature selection pipelines. Use of highest-ranking feature subsets from the training dataset yielded similar prediction in the testing dataset with AUC=0.74, sensitivity/specificity 0.66/0.70. Different algorithms showed different high-ranking features, but overall multiple data domains were represented among the highest-ranking predictors selected by each algorithm.

**Conclusions:** These findings provide evidence for feasibility of predicting youth SA using data collected at a single timepoint early in life in a diverse cohort. Results encourage incorporation of multiple data types including neurocognitive and geocoded environmental measures in machine learning SA prediction pipelines.

## Introduction

Suicide is the second leading cause of death in the US in adolescence, with rates of pediatric suicide (1) and suicide attempt (SA) rising in recent years (2), especially in Black youth (3, 4) . However, clinicians’ capacity to predict suicidal behavior is limited (5). Development of tools that can enhance SA risk stratification among youth addresses a major clinical gap that can improve youth suicide prevention (6). Suicidal behavior is a complex behavior associated with several biological and environmental aspects (7), and factors linked to suicidal behavior have been extensively studied at the individual- (e.g., psychological, cognitive) (8) and population-level (9). From a developmental perspective, data suggest that diverse risk factors come into play at different stages of life and suicide risk is a cumulative result of predisposing and precipitating risk factors (and their complex interactions) (8) over the life course (10). Scarce data exist regarding risk factors for suicidal behavior in childhood and early adolescence (11).

One strategy to address the challenge of SA prediction is using machine learning (ML) algorithms (12, 13). Research is now in the phase of determining the optimal way of leveraging such prediction algorithms into translational tools (14). A few promising studies suggest that ML predictive modeling can yield predictions of clinical relevance (15), mostly relying on the electronic health record [EHR] (16). Predicting SA in youth based on EHR data may be challenging, because the information contained in the average youth’s EHR is substantially less extensive than that contained in the typical adult’s EHR. Indeed, optimization of SA prediction will likely require use of more comprehensive predictor sets (17). Therefore, identification of predictors that can be added to the EHR and facilitate youth SA prediction is critical. Moreover, recent research indicates that SA prediction algorithms underperform in Black individuals (18). To address the challenge of Black youth suicide, research is needed on the predictive performance of SA algorithms that rely on data from diverse youth populations.

In the current study, we leveraged multimodal data (clinical, neurocognitive, environmental) in a diverse youth sample from the Philadelphia Neurodevelopmental Cohort (PNC, *T1*, collected between 2009-2011) (19) to predict self-report of a SA as documented in their EHR 1-10 years later in pediatric primary care (*T2a*) or emergency department (*T2b*) settings. We aimed to (i) test whether data collected in early-adolescence can contribute to the prediction of youth SA reported in mid/late adolescence; (ii) evaluate if different data domains, or combinations of selected predictors, will outperform prediction based on data from a single data domain; and (iii) evaluate performance of SA predictive algorithms when tested on an independent different clinical population than that used to train the algorithm.

## Materials and Methods

### Participants

We included youth who participated in the PNC study. The PNC sample (N=9,498, age range 8-21) was ascertained through the Children’s Hospital of Philadelphia (CHOP) pediatric care network between 2009-2011 (19). It included children in stable health, proficient in English, and physically and cognitively capable of participating in an interview and performing computerized neurocognitive testing. Notably, participants were not recruited from psychiatric clinics, and the PNC sample is not enriched for individuals who seek psychiatric help. Written assent and parental permission were obtained from all participants aged <18. University of Pennsylvania and CHOP’s Institutional Review Boards approved all procedures.

Here we included PNC participants for whom we identified an indicator in their EHR that they completed suicide attempt screening (N=1,419, Mean age 11.1 years, 53.3% Female, 51.4% Black race). The EHR included two different screening tools: the Patient Health Questionnaire-Modified for Teens (PHQ-9-M, available for n=922), a screening tool for depression and suicidal behaviors conducted routinely at CHOP’s well-child visits in adolescents ages 12 and older (20); Behavioral Health Screening–Emergency Department (BHS-ED, available for n=497), a mental health screening tool implemented at CHOP emergency department for youth ages 14 and older (21). **Table 1** details demographic characteristics of the study population.

**Table 1.** Sample Characteristics.

|  | **Overall**<br>**(N=1,419)** |  | **Training dataset**<br>**(n=922)** |  | **Testing dataset**<br>**(n=497)** |  | **P-value** |
| --- | --- | --- | --- | --- | --- | --- | --- |
|  | **N / mean** | **% / SD** | **N / mean** | **% / SD** | **N / mean** | **% / SD** |  |
| **Age at T1 (years)** | 11.08 | 2.2 | 10.46 | 1.71 | 12.24 | 2.52 | <0.001 |
| **Age at T2 (years)** | 16.67 | 1.85 | 16.24 | 1.63 | 17.47 | 1.96 | <0.001 |
| **Sex = Female** | 757 | 53.3 | 439 | 47.6 | 318 | 64 | <0.001 |
| **Race = White** | 522 | 36.8 | 369 | 40 | 153 | 30.8 | 0.001 |
| **Race = Black** | 729 | 51.4 | 449 | 48.7 | 280 | 56.3 | 0.007 |
| **Ethnicity = Hispanic/Latino** | 98 | 6.9 | 58 | 6.3 | 40 | 8 | 0.256 |
| **Average Parent Education** | 13.95 | 2.35 | 14.28 | 2.37 | 13.33 | 2.18 | <0.001 |
| **Parents Separated or Divorced** | 272 | 19.6 | 154 | 16.9 | 118 | 24.7 | 0.001 |
| **Suicide ideation at T1** | 72 | 512 | 38 | 4.1 | 34 | 7.1 | 0.026 |
| **Lifetime Suicide Attempt at T2** | 90 | 6.3 | 49 | 5.3 | 41 | 8.2 | 0.04 |

Note. T1 = first meeting with children, baseline; T2 = second meeting; Lifetime Suicide Attempt questions at PHQ-9-M: Have you EVER, in your WHOLE LIFE, tried to kill yourself or made a suicide attempt?; Lifetime Suicide Attempt questions at BHS-ED: Have you ever tried to kill yourself?

### Baseline PNC assessment (T1, mean age 11.1 years, conducted 2009-2011)

Participants underwent deep phenotyping that included an in-person clinical interview (for participants older than 10), and interview with the primary caregiver (for participants aged 8-17). Thus, ages 8-10 received only caregiver interviews and ages 11-17 received both caregiver and self-report. Interviews were conducted by trained and supervised assessors using a structured screening interview, based on Kiddie Schedule for Affective Disorders and Schizophrenia (K-SADS); and first-degree family history of psychiatric conditions was also recorded (19). All participants also underwent neurocognitive assessment with the Penn Computerized Neurocognitive Battery (22).

### Longitudinal assessment regarding history of SA

#### Community general pediatric clinic SA screening (T2a, mean age 16.2 years, conducted 2015-2018)

The CHOP network employs routine screening for depression and suicide risk using the PHQ-9-M, which includes nine core items regarding past 2-week depression, and two supplemental items assessing suicide risk (20). Of the 922 study participants who completed PHQ-9-M, n=884 (95.9%) completed their screening in CHOP pediatric clinics as part of a well visit. The remaining participants (<5%) completed the PHQ-9-M as part of routine follow up visit in one of CHOP’s specialty clinics (e.g., pulmonary, endocrinology, immunology, neurology).

#### Pediatric emergency department SA screening (T2b, mean age 17.5 years, conducted 2012-2019)

The BHS-ED is administered during clinical visits at CHOP emergency department. Adolescents without acute or critical injuries or illness are offered to complete the survey by the nursing staff (21). We identified 669 PNC participants who completed BHS-ED assessment at least one year after PNC assessment. Of these participants, 172 also completed the PHQ-9-M (i.e., were included in the training dataset) and were therefore excluded from the BHS-ED sample to allow separation between the training and the testing datasets.

### Measures

#### Outcomes

Endorsement of SA on the PHQ-9-M (“Have you ever, in your whole life, tried to kill yourself or made a suicide attempt?”) or on the BHS-ED (“Have you ever tried to kill yourself?”).

#### Predictors (i.e., features)

Predictors were derived from the PNC assessment (T1) and included a total of 193 data features. A full list of features is detailed in **Supplementary Table 1** and in **Supplemental eMethods**. Features were derived from 5 data domains including:

1. *Demographics* (7 features)
2. Clinical symptoms (131 features from PNC clinical assessment).
3. Past traumatic experiences (8 features) that were assessed as part of the PNC clinical interview (19).
4. Neurocognitive performance metrics (accuracy and speed, 26 features), which were previously associated with suicidal ideation in the PNC cohort (23).
5. Familial factors (7 features) including family history of psychiatric disorders, parental education and whether the parents are separated.
6. Geocoded neighborhood (census block-level) environment measures (14 features) that were obtained by linking geocoded participants’ addresses to census data (24), which were previously linked to suicidal ideation in PNC (25).

### Experimental design

First (**Step 1**), we examined performance of models using data collected at T1 to predict endorsement of SA at a later pediatric well-visit in the PHQ-9-M screening (T2a). We compared performance of a model trained on the entire dataset (193 features) to models that were trained on data from one of the six data domains.

Thereafter (**Step 2**), we ran three feature selection algorithms (Lasso, Relief, Random Forest) to identify specific features’ relevance to the prediction. We then tested whether a model that uses a subset of selected features yielded better prediction performance than a model relying on all 193 features.

Lastly (**Step 3**), we validated the models from step 2 with a different longitudinal dataset from a different clinical setting to predict endorsement of SA in an independent sample (T2b). **Figure 1** presents a schematic visual of the prediction pipeline.

**Figure 1.**
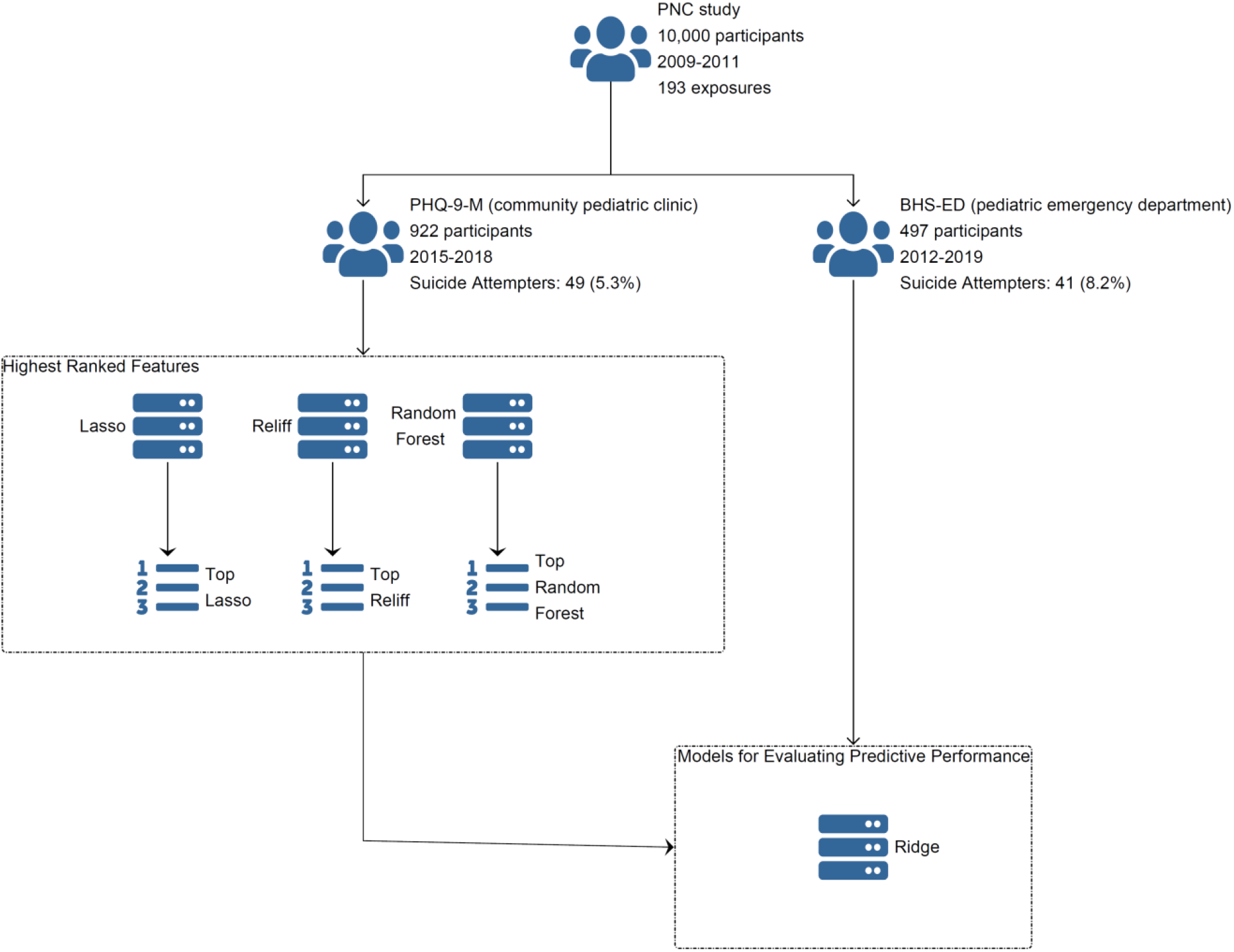
Schematic visual of the prediction pipeline.

### Statistical Analyses

#### Evaluation of model performance using entire datasets and specific data domains

We used area under the receiver operator characteristic curve (AUC) as the main metric to evaluate the prediction obtained with Ridge regression. Ridge is a form of regularized regression that assesses a “penalty” for the collinearity among the predictors in the model (26). Ridge regression was implemented 10,000 times using resampling (i.e., 10,000 “runs”). At the beginning of each run, the sample was divided randomly to 75% training set and 25% testing set.

#### Feature selection procedure and predictive performance of models using the selected features

We used three algorithms that allow identification of feature importance: Lasso regression (27), Random Forest (28), and Relief (29). These algorithms use different methods to ensure multiple-variable characteristics are considered in the feature selection process —e.g., besides main effects, does the variable have a nonlinear relationship with the outcome, does the variable interact with other variables in determining the outcome, and more. Further description of the feature selection algorithms and the rationale to choose them is detailed in **Supplementary eMethods**.

We used a 10-folds cross validation to run the algorithms above. The algorithms trained on 90% of the sample in each of the 10 folds. We therefore obtained three subsets of highest-ranking features (one using each algorithm). Afterwards, we trained Ridge on the same 90% of the sample but using the three subsets of the selected features. To each subset, we added the demographic features (age, sex, race) and duration between-visits. We then tested the prediction performance of these models using the remaining 10% of the sample. We used AUC, sensitivity, specificity, and positive and negative predictive values as performance metrics.

#### Predictive performance of models on an independent testing dataset

In our last step, we trained Ridge on the training dataset (from community pediatric clinic) using four sets of features. Three were feature subsets obtained from the feature selection models, Lasso, Relief and Random Forest. The fourth set included the entire feature list. We then tested the models’ performance using the independent testing dataset of youth screened for suicide attempt at the emergency department.

*Handling of missing data*

Missing data were imputed using two methods: maximum likelihood via expectation-maximization using the Amelia [II] package (30), and random forest using the missForest package (31). We previously used maximum likelihood imputation of some of these data, and to be consistent with previous publications, we have left that data imputed as originally released (24). However, more recent work has shown that random forest imputation is often superior to maximum likelihood (32), and we therefore imputed the remaining (newer) data (missing = 1%) using random forest (missForest). Features with more than 10% missing were removed from the analysis.

#### Sensitivity Analyses

We conducted two types of sensitivity analyses.

1. We reran all the steps without the clinical predictor “have you ever thought about killing yourself?”, to address the possibility that algorithm performance was mostly influenced by this feature.
2. To address the potential bias that could be caused by data imputation, we reran the models following listwise deletion of missing data.

## Results

### Predictive performance of youth SA using multimodal data and specific data domains

Inclusion of all 193 features showed better prediction (higher AUC) than models relying on specific data domains, with AUCs of 0.72 (supplementary **Figure 1**). Comparison across data domains indicated that the “clinical” domain yielded the highest predictive performance with AUC of 0.68. Other domains showed inferior performance, in descending order: “Family-level characteristics”, AUC=0.65; “demographics” and “geocoded neighborhood environmental measures”, for both AUC=0.60: “Neurocognitive”, AUC=0.56; and “Trauma exposure”, AUC=0.46.

### Predictive performance of youth SA using selected high-ranking features

We then tested whether subsets of selected features can yield better predictive performance then using all features. For that, we trained three feature selection algorithms (Lasso, Relief and Random Forest) in a 10-fold cross validation framework to select features relevant for the prediction. This allowed generation of three subsets of selected features (one from each algorithm) that we later used to predict SA using Ridge.

Models relying on the high-ranking feature subsets achieved better prediction performance than models that included all the 193 features together (**Table 2**). The greatest AUC of 0.75 was achieved using two subsets, the highest-ranking features chosen by Relief and Random Forest. These two models also achieved a balance between sensitivity/specificity with 0.76/0.77 and 0.80/0.74, respectively. The decision-rule for the cutoff was the closest point on the ROC Curve to (0,1), which usually optimizes the sensitivity/specificity tradeoff. These two models achieved Negative Predictive Value (NPV) and Positive Predictive Value (PPV) of 0.98/0.18 and 0.98/0.19 respectively **(Table 2)**.

**Table 2.**
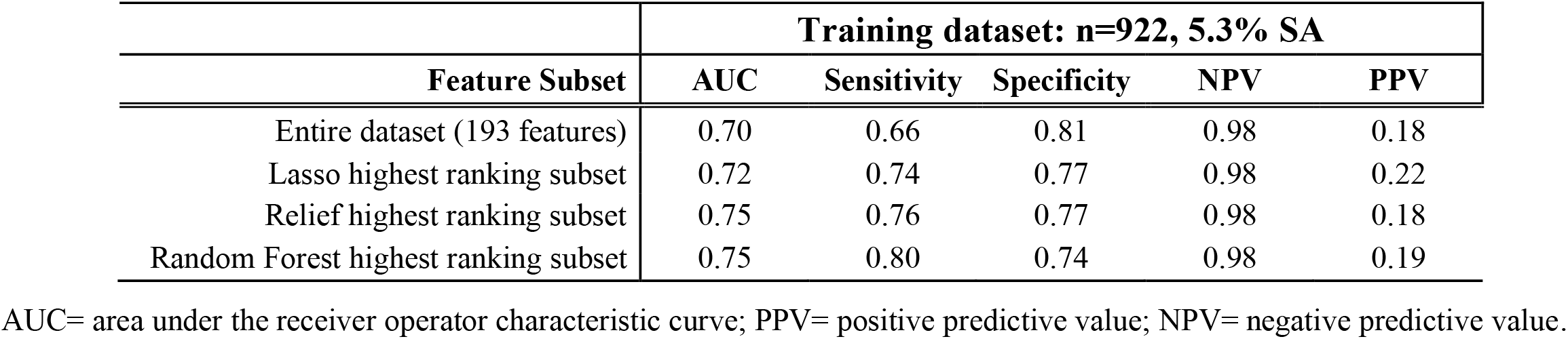
Predictive performance of suicide attempts in general community pediatric settings using different feature subsets.

### Characteristics of high-ranking features

To determine which features were most commonly selected by the three algorithms we averaged the number of times each feature was selected across the three algorithms, creating a ranked list. The highest-ranked predictor was the clinical feature from PNC evaluation (T1) “Have you ever thought about killing yourself?”. The next highest-ranking four features included two geocoded neighborhood environment features: median family income and percent of vacant lots in the neighborhood; and two neurocognitive features: speed of performance on an emotion intensity differentiation test and accuracy of performance on a test of recall memory for human faces. The ten highest-ranking features were from three different data domains: 2 clinical, 4 neurocognitive and 4 geocoded neighborhood environment features. **Table 3** presents the twenty highest-ranking features. For the full ranked list of all 193 features in each of the three feature selection algorithms see **Supplementary Table 2**.

**Table 3.** The 20 highest ranked features obtained when averaging feature importance ranking among three algorithms.

| Rank | Domain | Feature recorded at T1 |
| --- | --- | --- |
| 1 | Clinical | Have you ever thought about killing yourself? |
| 2 | Neurocognitive | Social cognition: emotion intensity discrimination (speed) |
| 3 | Neighborhood | Median family income |
| 4 | Neurocognitive | Memory for Faces (accuracy) |
| 5 | Neighborhood | Percent vacant lots |
| 6 | Neighborhood | Percent white |
| 7 | Neurocognitive | Memory for Faces (speed) |
| 8 | Neurocognitive | Verbal Memory (accuracy) |
| 9 | Neighborhood | Percent Female |
| 10 | Clinical | Do people ever seem to have difficulty understanding you? |
| 11 | Demographics | Sex |
| 12 | Neurocognitive | Complex reasoning: language / verbal Reasoning (speed) |
| 13 | Neighborhood | Percent in Poverty |
| 14 | Neighborhood | Percent Married |
| 15 | Family Environment | Parents Separated or Divorce |
| 16 | Clinical | I think I might feel like my mind is "playing tricks" on me. |
| 17 | Neurocognitive | Social cognition: age differentiation test (accuracy) |
| 18 | Neurocognitive | Executive Function, Attention (speed) |
| 19 | Neighborhood | Percent Non-Family Households |
| 20 | Clinical | Have there been times when you kept talking a lot, couldn't stop talking, talked faster than usual, had thoughts faster than usual, or had so many ideas in your head that you could hardly keep track of them? |

### Generation of selected high-ranking feature subsets

We created three “lean” feature subsets using high-ranked features. Three subsets were created based on the results of the three feature-selection algorithms. For each algorithm, the subsets included predictors that were selected in at least half of the folds (more than 5 times), resulting in 7 features for Lasso, 60 for Relief and 45 for Random Forest. The feature subsets included predictors from 5 different data domains when using Relief and Random Forest (clinical, demographic, neurocognitive, family, neighborhood), and included 2 domains when using Lasso (clinical and demographic) (**Figure 2**).

**Figure 2.**
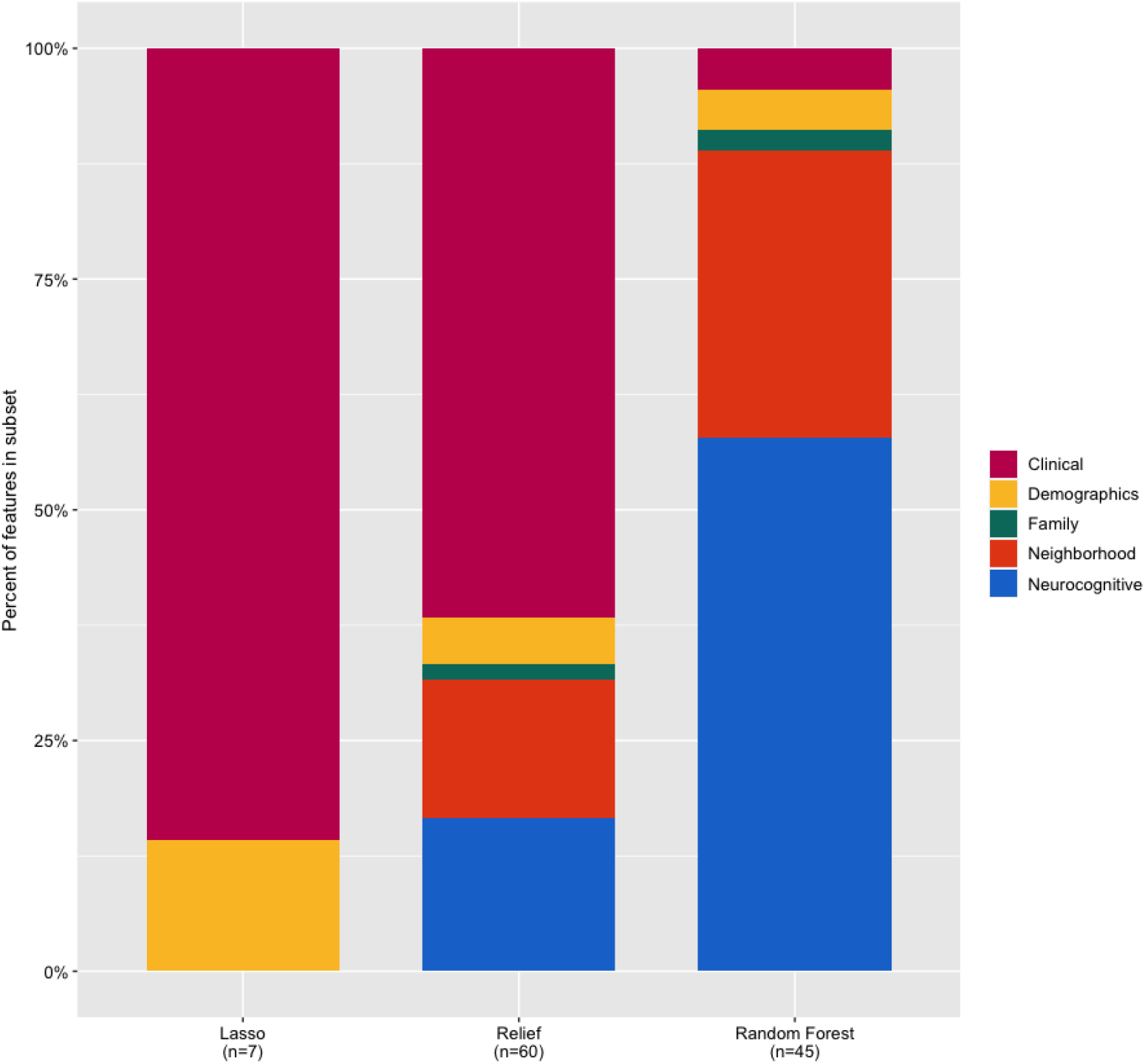
Distribution of the different data domains in each of the three subset datasets composed of top-ranked features.

### Testing youth SA prediction algorithms on an independent clinical sample

To assess generalizability of findings to a different clinical youth population, we tested the SA predictive performance of the algorithms, which were trained on data from general community pediatric clinics, in an independent longitudinal sample of youth presenting to a pediatric emergency department. We tested prediction performance of algorithms using 4 feature sets: either the entire dataset or using the three high-ranked feature subsets described above.

The best performing algorithm in the independent replication sample yielded an AUC of 0.74 and was achieved by the model using the Lasso derived feature subset (sensitivity = 0.66, specificity= 0.70 NPV= 0.96, PPV = 0.17). All other models achieved AUCs between 0.62 and 0.70 **(Table 4)**.

**Table 4.** Predictive performance of suicide attempts in an independent testing dataset of pediatric emergency department settings using different feature subsets identified in a training dataset from general community pediatric settings.

|  | **Testing dataset, n=497, 8.2% SA** |  |  |  |  |
| --- | --- | --- | --- | --- | --- |
| **Feature Subset** | **AUC** | **Sensitivity** | **Specificity** | **NPV** | **PPV** |
| Entire dataset (193 features) | 0.70 | 0.61 | 0.75 | 0.96 | 0.18 |
| Lasso top 7 feature subset<sup>a</sup> | 0.74 | 0.66 | 0.70 | 0.96 | 0.17 |
| Relief top 60 feature subset<sup>a</sup> | 0.70 | 0.66 | 0.68 | 0.96 | 0.16 |
| Random Forest top 45 feature subset<sup>a</sup> | 0.63 | 0.61 | 0.57 | 0.94 | 0.11 |

<sup>a</sup> Subsets included only features that were included in at least 5 of the 10-fold validation feature subsets selected in the training dataset.
AUC= area under the receiver operator characteristic curve; PPV= positive predictive value; NPV= negative predictive value.

### Sensitivity Analyses

To address the possibility that algorithm performance was mostly influenced by the feature of endorsing suicidal ideation at baseline PNC assessment, we reran analyses excluding this feature. The prediction was slightly lower with AUC=0.70 for the best performing model vs. 0.75 obtained when including the suicidal ideation feature. Full models’ statistics are in **Supplementary Table 3**.

To address the possibility that results were biased by imputation, we used listwise deletion and found that the performance of predictive models was similar, with AUC of 0.72 for the best performing model vs. 0.75 in the imputed dataset used in main analyses. Full models’ statistics are in **Supplementary Table 4**).

## Discussion

We provide evidence that data collected in a single timepoint in pre- or early adolescence can help predict self-report of SA by mid-late adolescence in a community pediatric setting. The prediction achieved good discrimination (AUC=0.75) that was replicated in an independent dataset. This performance is comparable to previous work that used longitudinal EHR data to predict SA in older youth (33, 34), and greater than a previous study using prenatal and postnatal environment data to predict adolescent SA (35). The application of our algorithm derived from suicide attempt screening data in community settings to an independent sample from pediatric emergency department setting bears promise for future clinical translation of our approach combining multiple data types to predict youth SA. Our findings suggest that multiple data types (clinical, neurocognitive and geocoded neighborhood environment data) can contribute to adolescent SA prediction.

This study adds key insights to the field as it navigates through the challenges of incorporating machine learning strategies to optimize youth suicide prevention (36). First, we provide evidence that machine learning algorithms can help predict SA in a diverse population with >50% Black youth, a subgroup showing a concerning, upward trend in suicide in recent years (3, 4). This is important in light of evidence suggesting that the risk factors for Black youth SA are distinct (37). Second, we show that a single assessment in early age (pre/early adolescence), a developmental epoch with limited data on relevant suicide risk factors (11), can generate a reliable SA prediction by the time youth reach an age when SA risk peaks (mid-late adolescence). Third, by comparing three different algorithms and selecting different features subsets, we found that (i) each algorithm makes use of different predictors from various domains; and (ii) the top performing models rely on predictors selected from different data domains. This may suggest that multiple data domains can prove useful in optimizing youth SA predictions and that no data domain can be simply “ruled out” as irrelevant. Our work provides empirical support for the notion that clinically useful predictive algorithms will benefit from diverse sets of predictors (17).

A key consideration when interpreting our results regarding feature importance to the prediction is to refrain from causal inference (17). Research should carefully make the distinction between improving the prediction capability of clinicians and understanding the causal pathways leading youth to attempt suicide (38). Our study addresses the former approach. However, it is reassuring to see that suicidal ideation at baseline was the highest-ranked predictor. We suggest that when the time comes for clinical translation, identification of strong clinical indicators at the top of the predictor list would bolster confidence in clinicians using prediction algorithms, which is critical for their implementation (39). We also emphasize that one should avoid interpretation of the cognitive predictors as causal because this was not the study aim. Rather, we suggest that our results provide proof-of-concept for the incorporation of neurocognitive tests (which can be taken as early as age eight (19) in ML algorithms predicting SA. Lastly, it is notable trauma exposure did not emerge as a high-ranking predictor in any of the models. While this finding may be surprising because history of early life adversity is an established risk factor for youth SA (11), it is consistent with a recent study using clinical data and stress exposures measure to predict SA in German youth, where trauma exposure was ranked relatively low (bottom half) among 16 predictors from clinical and sociodemographic domains using four different algorithms (40).

Limitations of this study should be noted. First, ML algorithms are not intended to allow causal interpretation, hence it is difficult to understand how the highest-ranking predictors contribute to SA prediction. Understanding the individual contribution of the predictors to SA is outside the scope of this work and will likely require utilization of different approaches geared toward interpretability and causal inference. Second, this study included participants ascertained in a single site, and generalizability outside this site is challenging. Nonetheless, the fact that results replicated in an independent sample at different clinical setting is encouraging. Relatedly, our sample size of 1,419 is sufficient for providing proof-of-concept but will require replication in larger datasets prior to implementation of predictive algorithms in clinical settings. Fourth, the predicted outcome was report of SA 5-6 years (on average) after the baseline assessment. We cannot rule out the possibility that SA occurred prior to baseline assessment. However, since the participants were very young at baseline (mean age 11), it is unlikely that the SA occurred prior to baseline assessment. Notably, even in the case that it did occur before the baseline assessment, our results are still clinically meaningful in their capacity to discriminate suicide attempters from controls using data collected at an early age.

In conclusion, we describe good predictive capacity of ML algorithms that maintained good performance when tested in an independent dataset from a different clinical population. The predictive algorithms used predictors from various data domains, some of which are not usually found in youth EHR. The young age of the sample and the inclusion of >50% Black youth address a major clinical gap and provides evidence for feasibility of predicting SA with clinical, neurocognitive and geocoded neighborhood environment’s data early in or even before adolescence. Findings point to types of data that can be easily collected and integrated in youth EHR (especially geocoded measures) to optimize SA prediction. Future works are needed to test implementation and acceptability of such tools by clinicians and patients.

## Data Availability

Data used for prediction of suicide attempts is available in deidentified format at dbGAP. Data regarding suicide attempts was derived from medical records of patients and cannot be shared. The authors are open to collaborations upon request while adhering to privacy policies.

https://www.ncbi.nlm.nih.gov/projects/gap/cgi-bin/study.cgi?study_id=phs000607.v3.p2

## Acknowledgments

This study was supported by the National Institute of Mental Health grants R21MH123916, K23MH120437 and the Lifespan Brain Institute of Children’s Hospital of Philadelphia and Penn Medicine, University of Pennsylvania.

## Notes

**Conflict of Interest Disclosures:** Dr Barzilay serves on the scientific boards and receives consulting fees from ‘Taliaz Health’ and ‘Zynerba Pharmaceuticals’, and holds equity in ‘Taliaz Health’, with no conflict of interest relevant to this work. All other authors have no conflicts of interest to disclose.

### Competing Interest Statement

Dr Barzilay serves on the scientific boards and receives consulting fees from Taliaz Health and Zynerba Pharmaceuticals, and holds equity in Taliaz Health, with no conflict of interest relevant to this work. All other authors have no conflicts of interest to disclose.

### Author Declarations

University of Pennsylvania and CHOP Institutional Review Boards approved all procedures.

